# Effect of Injectable Dual and Single Agonist Glucagon-Like Peptide-1 Based Therapy on Inflammatory Bowel Disease Activity Among Patients with Obesity

**DOI:** 10.1101/2025.11.13.25340005

**Authors:** Jake Levine, Yao An Lee, Angela Pham, Jingchuan Guo, Hao Dai, Rotana M Radwan, Jiang Bian, Aleksey Novikov, Amy Sheer

## Abstract

**Background and Aims:** Obesity is a risk factor for relapsing inflammatory bowel disease. GLP-1 and dual GLP-1/GIP agonists may offer superior weight loss; however, their effect on inflammatory bowel disease remains unknown. In this study, we assessed outcomes in patients with IBD and obesity using these medications.

**Methods:** A retrospective, propensity-matched analysis was conducted using data from The OneFlorida+ network. We evaluated patients with IBD and obesity prescribed GLP-1-based therapy (liraglutide, semaglutide, or tirzepatide) versus those without. 562 patients prescribed GLP-1-based therapies were matched 1:1 to controls based on demographics and comorbidities.

**Results:** No differences in GLP-1 users versus non-users with respect to all-cause hospitalization, IBD-hospitalization, IBD-related surgery, or pancreatitis were seen. Use of semaglutide reduced the risk of IBD-related surgery (HR 0.33, CI 0.13-0.83). Among 89 tirzepatide users, none required IBD-related surgery versus 2 matched controls. Black GLP-1 users had an increased risk of all-cause hospitalizations (HR 1.59, CI 1.11–2.29) but not IBD-related hospitalization or IBD-related surgery. Steroid use was comparable between groups. GLP-1 use significantly reduced serum CRP.

**Conclusions:** Semaglutide appeared to lower the risk of IBD-related surgery. Black patients using GLP-1 therapy had an increased risk of all-cause hospitalization. Tirzepatide showed no significant difference regarding primary endpoints; however, the sample size was small. Overall, all GLP-1-based therapies were well tolerated among obese patients with IBD. Larger studies are warranted to understand the therapeutic potential of GLP-1-based treatment in this patient population.

**Graphical Abstract:** 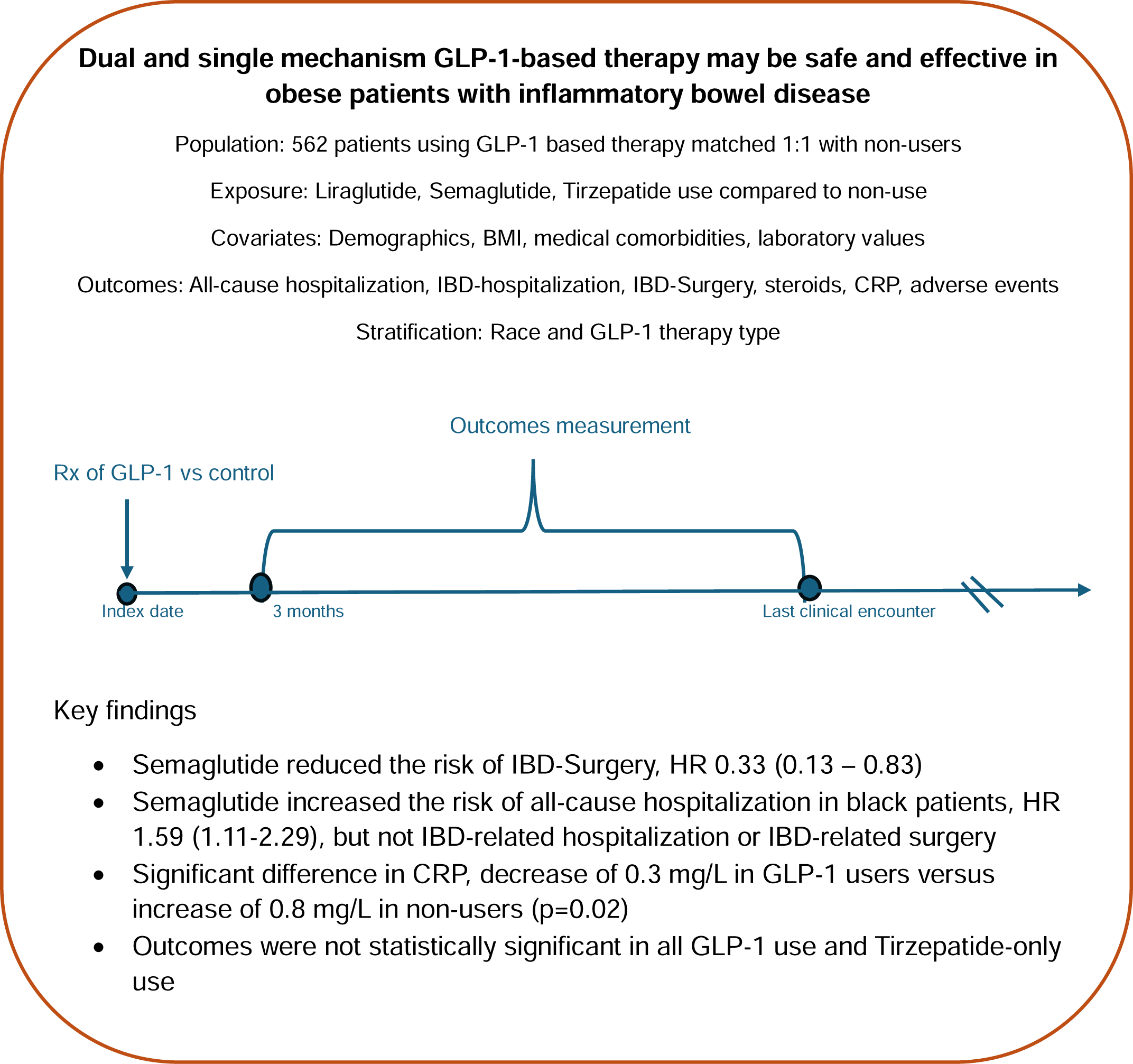

## Introduction

Patients with inflammatory bowel disease (IBD), a condition characterized by chronic inflammation in the GI tract, may suffer from a myriad of comorbidities secondary to their chronic disease^1,2^. The role of systemic inflammation in IBD has been well described in the available literature ^2^. As a result of this systemic inflammation, patients may develop increasingly severe symptoms or suffer from extraintestinal manifestations, such as inflammatory dermatoses, arthropathies, or ocular complications, among others. There is ongoing research into how lifestyle modifications may modulate the level of systemic inflammation in this patient population, and therefore the disease course ^3^. Many factors, such as age, sex, tobacco and alcohol use, level of physical activity, and others, may modulate inflammation ^4^. There is growing interest in the effect of obesity on IBD activity. Obesity is known to induce adipose-dependent expression of inflammatory mediators, such as TNF-alpha and IL-6^5^. Increased obesity may increase the chance of losing response to drugs by altering the pharmacokinetics of biologic therapy and may negatively impact the IBD disease course ^5^. In fact, Yarur et al showed that increased adiposity may act as a sink for biologic therapy, reducing clinical response ^6^.

Evidence is mounting that treating obesity may modify inflammatory pathways that underpin autoimmune diseases like IBD^7^. In fact, Neto et al showed that in a matched case series of patients undergoing bariatric surgery, the surgery cohort was less likely to require rescue steroids or require IBD-related surgery ^8^. With the advent of non-surgical weight loss treatments, there is increased attention to how these therapies impact IBD. Incretin therapies such as Glucagon-Like Peptide-1 (GLP-1) agonists like semaglutide have been studied to determine their effect on IBD activity ^9^. Despite initial concerns about its use in IBD, primarily due to gastrointestinal side effects like diarrhea and constipation, early studies are showing safety and reduction in IBD-related complications like intestinal obstruction^9^.

Novel injectable incretin therapies with dual GLP-1 and Glucose-dependent Insulinotropic Polypeptide (GIP) activity, such as the GLP-1/GIP agonist Tirzepatide, are now FDA approved for use in the treatment of obesity ^10^. However, there is a paucity of evidence regarding the safety of dual GLP-1/GIP agonists in the population of obese patients with IBD, and this study aims to add to the body of evidence on use in this patient population.

## Materials and Methods

### Data Source

Patient data was obtained from The OneFlorida+ clinical research network, which is part of the National Patient-Centered Clinical Research Network (PCORnet). The dataset spans from 2014 through 2024 and includes comprehensive, longitudinal electronic health records (EHRs) for approximately 21 million individuals across Florida, Georgia, and Alabama. OneFlorida+ integrates data from 14 partnering healthcare systems and is formatted according to the PCORnet Common Data Model (CDM).^11^ The dataset, designated as a HIPAA-limited set, includes key temporal and geographic identifiers, such as dates and ZIP codes, and provides robust clinical and demographic information.

### Study Design and Study Population

We conducted a retrospective cohort study with an intention-to-treat analysis to evaluate the association between GLP-1 therapies and outcomes in adults with IBD and Obesity. The study protocol was approved by the University of Florida Institutional Review Board prior to initiation, under IRB202300903.

The cohort included adults with IBD who met the eligibility criteria for anti-obesity medication, had a documented diagnosis of obesity (BMI ≥ 30 kg/m²) or a BMI in the range of 27.0–29.9 kg/m² with at least one weight-related comorbidity. Individuals with IBD were identified using a validated, EHR-based algorithm with diagnosis codes as detailed in Supplementary Table 1. The inclusion of patients using GLP-1 therapy versus non-users is outlined below.

### Covariates

Baseline covariates encompassed a range of demographic and clinical characteristics. Demographic variables included age, sex, race/ethnicity, smoking status, and insurance status. Clinical covariates included comorbid conditions (for example, cardiovascular disease, biliary disorders), biometric data (such as BMI, blood pressure), laboratory values (for example, HbA1c, GFR, lipids), and medications (such as aspirin, opioids, and statins). All Covariates are detailed in Supplemental File 1.

### Exposure of interest

The exposure of interest was the use of GLP-1-based therapies, liraglutide, semaglutide, or tirzepatide, identified using RxNorm codes and National Drug Codes (NDCs). The comparator group consisted of matched individuals who did not use GLP-1 therapies. The index date for GLP-1 users was defined as the date of the first GLP-1 prescription. The comparator group, defined as patients with IBD and obesity who did not initiate GLP-1 therapy during the study period, was constructed by aligning non-users with GLP-1 users within a ±1-week window of the initiation date, to ensure temporal comparability of treatment assignment. GLP-1 users were propensity matched 1:1 with non-users based on several clinical and demographic variables, as noted in Supplemental File 1, to control for confounders. Individuals were excluded if they were under 18 years of age at the index date, had not been diagnosed with IBD before the index date, or lacked any clinical encounters before the index date. Patients’ follow-up began at the index date. It continued until the earliest occurrence of one of the following censoring events: the outcome of interest, death, loss to follow-up (as indicated by the date of the last recorded clinical encounter), or the end of the study period (January 31, 2024).

### Study Outcomes

Primary outcomes included incidence of IBD surgery, all-cause hospitalization, and IBD-hospitalization. Secondary outcomes included risk of IBD surgery, all-cause hospitalization, and IBD hospitalization with respect to race and type of GLP-1-based therapy, change in C-reactive protein (CRP) from baseline to follow-up, steroid use, pancreatitis, and bowel obstruction. IBD surgeries were identified via CPT procedure codes outlined in supplementary table 1. IBD-hospitalization was defined as an admitting diagnosis of IBD or its subtypes based on ICD-10 diagnosis codes as noted in supplementary table 1.

### Statistical Analysis

To address potential confounding and enhance comparability between the two treatment groups, we implemented a time-dependent 1:1 propensity score matching strategy^13^. Propensity scores were derived from a multivariable logistic regression model incorporating a comprehensive set of baseline covariates. Matching was performed using a nearest-neighbor (KNN) algorithm without replacement and a caliper of 0.2, ensuring that each GLP-1 user was matched to a non-user with the closest propensity score within the specified threshold^14^. Covariate balance between matched pairs was evaluated using standardized mean differences (SMDs), with values below 0.1 considered acceptable balance between GLP-1 users and non-users. Furthermore, Cohen’s d statistic was used to assess the magnitude of difference in covariates before and after matching, providing additional assurance that confounding was adequately minimized in the final matched cohort used for outcome analyses^15^.

We calculated the incidence rate of study outcomes, and Cox proportional hazard regression models were used to estimate hazard ratios with 95% confidence intervals by comparing GLP-1 users with non-users.

## Results

### Study Population

The cohort included 1,124 patients with IBD and Obesity matched 1:1 and followed for a median of approximately 12 months. Among these at the index date, 475 had Crohn’s disease, 551 had ulcerative colitis, and 98 had indeterminate colitis. Age, sex, baseline BMI, and IBD subtype were comparable between groups (Table 1).

**Table 1:**
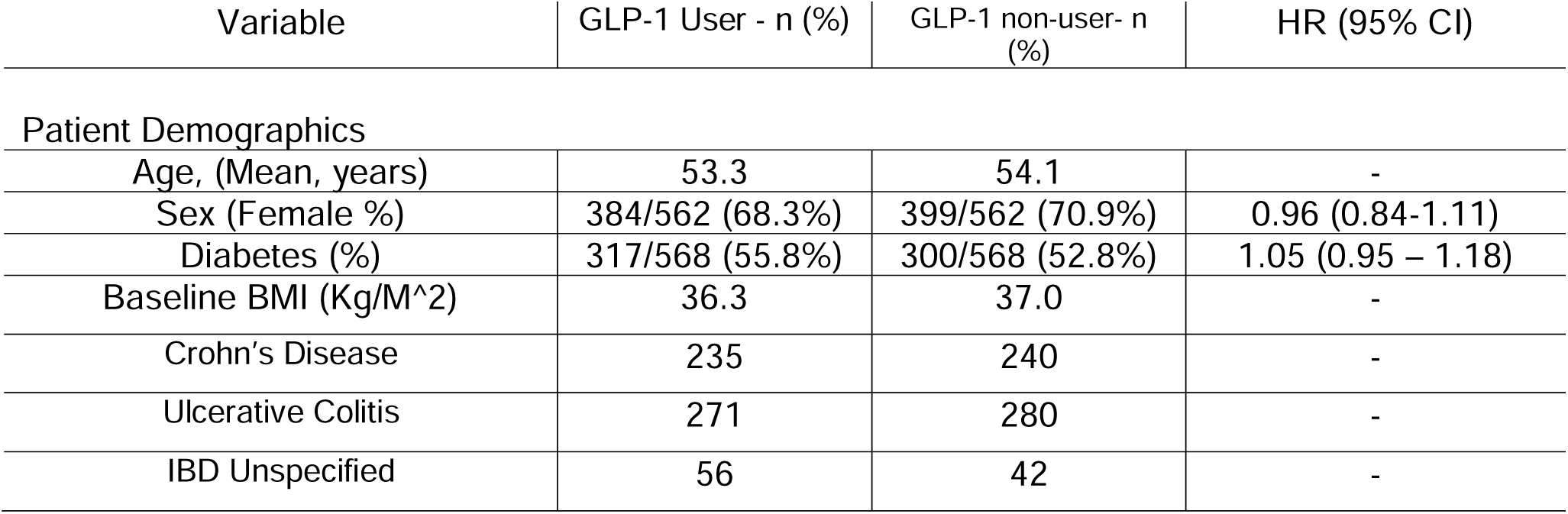
Description of Demographics and Statistical Analysis - HR= Hazard ratio; CI= Confidence interval; IBD=Inflammatory Bowel Disease; Sum of different GLP-1 agents is greater than 562, as some patients switched agents; Sum of self-identified race is less than 562, as some patients did not self-identify their race.

### Outcomes and Adverse Events

Among GLP-1-based therapy users versus non-users, there were no significant differences in all-cause hospitalization (HR 1.04, 0.85-1.28) or IBD-related hospitalization (HR 1.30, 0.90, 1.88). Furthermore, there was no significant difference in IBD-related surgery (HR 0.49, 0.21 – 1.17). While not statistically significant, zero patients required steroids in the GLP-1-based group, versus 5 in the non-user group. Rates of pancreatitis were low and similar between groups (HR 0.74, 0.37–1.47). Over a median follow-up of 9 months, GLP-1 users experienced a median decrease in CRP of 0.3 mg/L, whereas non-users had a median increase of 0.8 mg/L. These results were statistically significant (p = 0.02). Because 35.3% of patients had CRP levels, these changes could not be stratified further. These findings are summarized in Table 2.

**Table 2:**
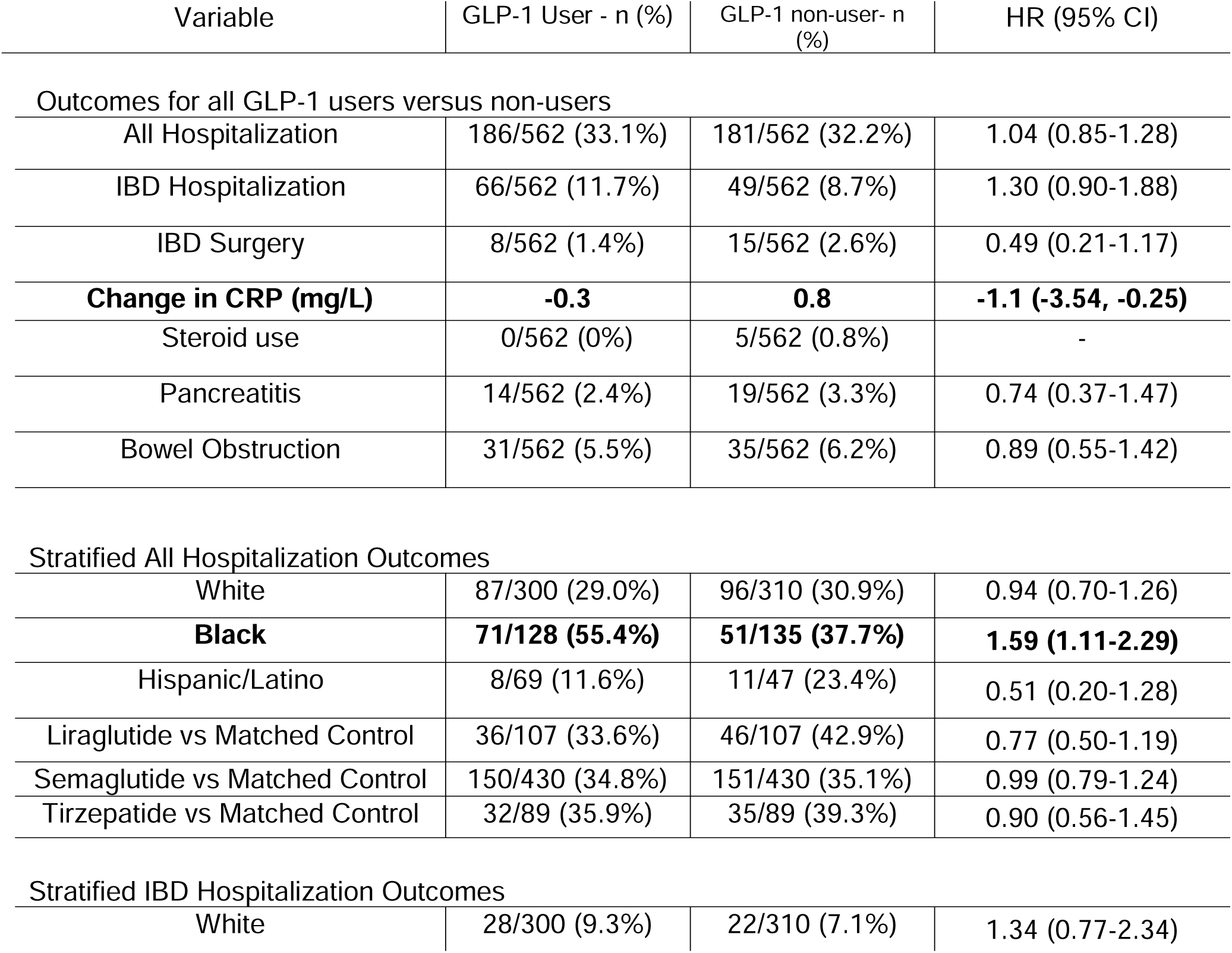
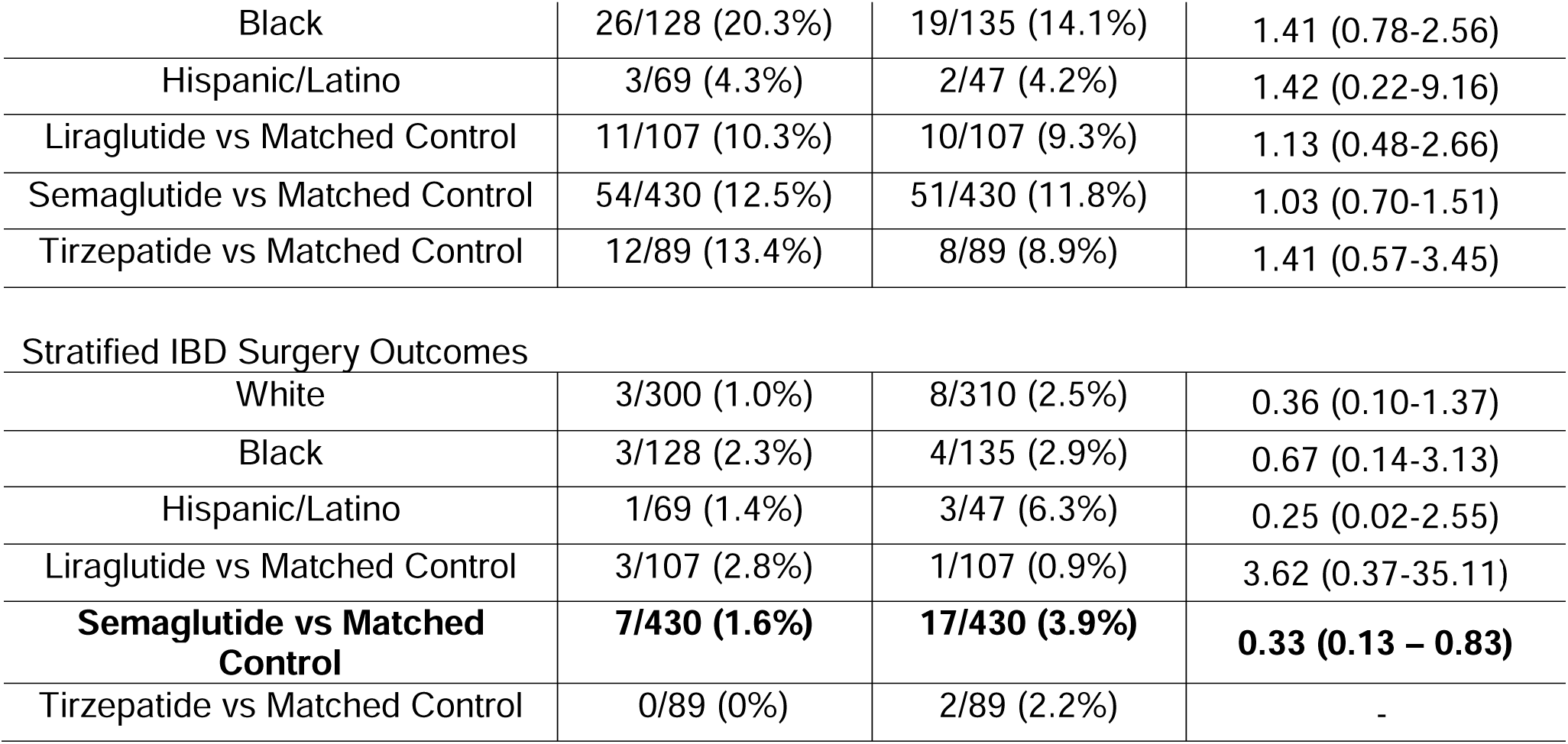
Primary and secondary outcomes.

### Subgroup Analysis

Outcomes were stratified by race and GLP-1-based drug in Table 2. Black patients in the GLP-1-based group were noted to have a higher risk of all-cause hospitalization (HR 1.59, 1.11-2.29), though they did not have a higher risk of IBD-related hospitalization (HR 1.41, 0.78-2.56). In addition, semaglutide use was associated with a lower risk of IBD-related surgery (HR 0.33, 0.13-0.83). It should be noted that tirzepatide use was not associated with a change in risk of the outcomes of interest. However, there were relatively fewer tirzepatide users as noted in Table 2.

## Discussion

Among all patients in this study with IBD and obesity, overall use of GLP-1-based therapies was not associated with improvement in all-cause hospitalization, IBD-specific hospitalization, or IBD-related surgery, nor was there evidence of harm. While a benefit in the outcomes of interest was not observed, there appears to be an indication that it is safe and well-tolerated, particularly in that there was no signal for pancreatitis or bowel obstructions, which are adverse events of concern in GLP-1-based therapy, especially in this population. 10 As expected, there were modest improvements in serum CRP, whereas CRP increased in those not using GLP-1 therapy.

Unexpectedly, there was an increased risk of all-cause hospitalization in black patients using GLP-1-based therapies compared to black patients not using them. It should be noted that there is literature showing that black patients using GLP-1-based therapies lose less weight on average than white patients, with one potential contributing factor being genetic ancestry-based differences in the *TCF7L2* gene, which modulates response to incretin-based therapies^16^. Lower weight loss or lack of incretin response may have had downstream effects that increase the risk of hospitalization. There is also well-described evidence that social determinants of health with respect to race may disproportionately affect black patients in ways difficult to ascertain in this study^17^. Larger studies should be conducted to see if this observation is reproduced.

In this study, semaglutide use alone was associated with reduced incidence of IBD-related surgery. CRP was also significantly lower among all GLP users compared to non-users. This confirms prior findings that have shown that GLP-1 therapies change the expression of many pro-inflammatory signals, such as epidermal growth factor (EGF), hepatocyte growth factor (HGF), IL-6, IL-1β, among others^18^. Such immunomodulatory effects may underlie the protective association observed with semaglutide in our cohort. This supports the idea that systemic inflammation improves with weight loss. This may potentially change the course of IBD and lower the risk of surgery. While we anticipated that tirzepatide, given its greater weight loss efficacy relative to semaglutide, might yield even more pronounced improvements in IBD outcomes, our analysis was limited by the relatively small number of tirzepatide users, which raised the possibility of underpowering and a type II error^19^. Nevertheless, tirzepatide appeared safe and well-tolerated in this patient population. Whether the benefits observed arise solely from weight loss or from the direct anti-inflammatory properties of GLP-1 signaling requires further study. Still, our findings contribute to growing evidence that metabolic therapies may influence immune-mediated disease pathways.

Pancreatitis is associated with IBD and can cause significant clinical problems^20^. In our study, the use of GLP-1 medications did not appear to increase the risk of pancreatitis in the population of patients with IBD. This is important to note because one of the most common reasons for withholding GLP1 agonists in clinical practice is their reported risk of pancreatitis^21^. Our data support prior data showing no risk of pancreatitis in patients on GLP-1 agonist therapy^22–24^.

These results naturally raise the question of whether GLP-1 or dual GLP/GIP agonists should be considered for patients with IBD who also have obesity. While our findings provide reassurance that these therapies are generally well tolerated and may confer secondary benefits beyond weight loss, they do not establish causality. They should not be interpreted as practice-changing at this stage. Clinicians should continue to follow guideline-based care, while recognizing that the use of GLP-1–based therapy for obesity in IBD patients may provide both metabolic and potentially disease-modifying benefits. Notably, emerging trials are investigating GLP-1 receptor agonists in IBD (e.g., NCT05196958, NCT06774079), underscoring the growing interest in combining metabolic modulators with immunosuppressive therapies in IBD.

This study had notable limitations. First, the study examined composite outcomes from a large dataset, so specific details, such as the reason for all-cause hospitalization and the exact type of IBD surgery, were not ascertained. Additionally, some patients switched between multiple GLP-1-based therapies, which were not accounted for in the analysis. In addition, surrogate markers for disease activity were used, such as hospitalization, steroid use, CRP, and IBD-related surgery. More proximal markers of disease, such as validated symptom scores, fecal calprotectin, changes in advanced IBD therapy, and endoscopic disease activity scores, were not assessed. This may lead to some difficulty in interpreting disease severity in this patient population. Additionally, the degree of weight change between these groups was not evaluated.

In summary, novel GLP-1-based therapies like tirzepatide appear safe and well-tolerated in the IBD population. As mentioned above, larger trials should be conducted to explore these clinical questions further. Attention should be paid to differential effects among racial and ethnic groups. Future prospective clinical trials are needed to evaluate GLP-1 and GLP/GIP agonists in patients with IBD and obesity, with careful attention to disease activity, inflammatory biomarkers, and health disparities. If confirmed, these therapies could represent a paradigm shift, where obesity treatment is leveraged not only for its metabolic advantages but also as an inflammation-modifying strategy in chronic immune-mediated diseases. Future studies should also examine whether particular IBD subtypes, treatment backgrounds, or patient demographics influence therapeutic response. Ultimately, integrating obesity management into IBD care may represent a promising, multifaceted approach to improving both metabolic and gastrointestinal outcomes.

**Figure 1:**
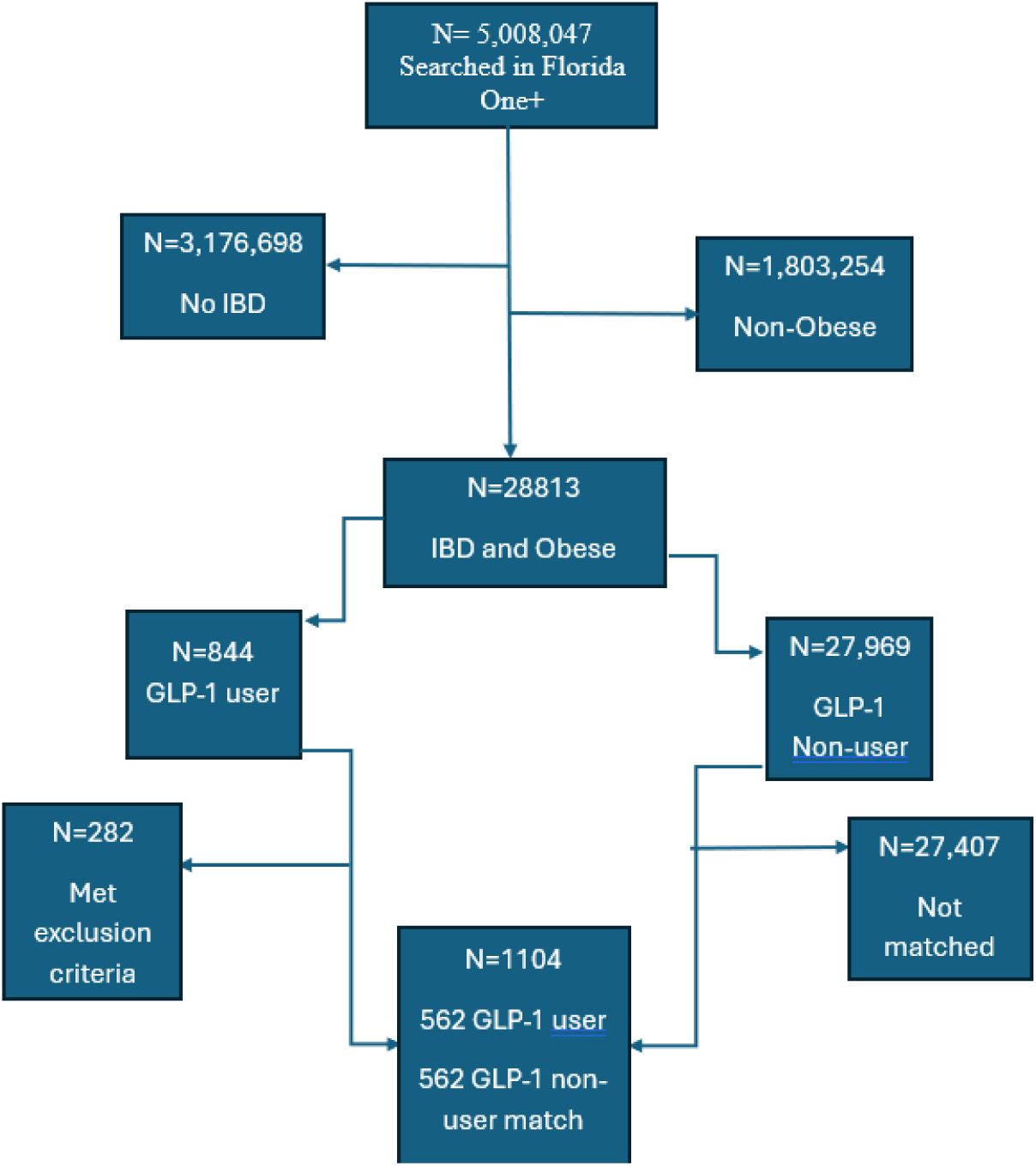
Flowchart for patient selection.

## Financial Conflicts of Interest

Angela Pham

- Abbvie Consultant

- Janssen Advisory Committee and Board Member

- Grant/Research Support from Pfizer Inc.

Amy Sheer Joy

- Consultant for PeerView

All other authors declare no financial conflicts of interest

## Acknowledgments

We would like to acknowledge the OneFlorida+ network database and the patients within it for making this study possible.

We would like to thank Dr. Johan Nordenstam for assisting in finding relevant CPT codes for IBD-related surgery.

The manuscript, including related data, figures, and tables, has not been previously published or submitted for publication elsewhere. This manuscript is not under consideration elsewhere.

## Authors Contributions

Jake Levine – Conceptual development of project, development of methods, conceptual development of data collection and outcomes, study design, drafting of manuscript, co-first author

Yao An Lee – development of methods, data extraction, data analysis, drafted the methods section of manuscript, manuscript editing, co-first author

Angela Pham – development of methods, study design, manuscript editing

Jingchuan Guo – development of methods, study design, data extraction, data analysis

Hao Dai - data extraction, data analysis

Rotana M Radwan - data extraction, data analysis, manuscript editing

Jiang Bian - data extraction, data analysis

Aleksey Novikov – development of methods, study design, manuscript editing

Amy Sheer – development of methods, study design, senior author, manuscript editing

## Supplementary Data

Included in separate files. Includes supplementary file 1 and supplementary table 1.

## Data Availability Statement

The data underlying this article cannot be shared publicly due to its use on other projects for which the respective investigators have not given permission for data sharing. Data will be shared on reasonable request to the corresponding author.

