## Supplemental Table for "Effect of Injectable Dual and Single Agonist Glucagon-Like Peptide-1 Based Therapy on Inflammatory Bowel Disease Activity Among Patients with Obesity"

| IBD Disease | ICD-10 |
|---|---|
| Crohn's Disease | K50, K50.0, K50.1, K50.8, K50.9 |
| Ulcerative Colitis | K51, K51.0, K51.2, K51.3, K51.5, K51.9, K52.3 |
| **Surgery Name** | **CPT** |
| Colon resection | 44140 |
| Small bowel resection | 44120 |
| Ileostomy | 44143 |
| Colostomy | 44144 |
| j-pouch creation | 44155 |
| Ileocolic resection | 44160 |
| Subtotal colectomy | 44150 |
| IPAA | 44157 |
| Strictureplasty | 44615 |
| Small Intestine and Colon | 43820, 43860, 43865, 44110, 44111, 44112, 44113, 44114, 44115, 44116, 44117, 44118, 44119, 44120, 44121, 44122, 44123, 44124, 44125, 44126, 44127, 44128, 44129, 44130, 44131, 44132, 44133, 44134, 44135, 44136, 44137, 44138, 44139, 44140, 44141, 44142, 44143, 44144, 44145, 44146, 44147, 44148, 44149, 44150, 44151, 44188, 44202, 44203, 44204, 44205, 44206, 44207, 44208, 44209, 44210, 44211, 44212, 44213, 44214, 44215, 44216, 44217, 44218, 44219, 44220, 44221, 44222, 44223, 44224, 44225, 44226, 44227, 44228, 44229, 44230, 44231, 44232, 44233, 44234, 44235, 44236, 44237, 44238, 44300, 44301, 44302, 44303, 44304, 44305, 44306, 44307, 44308, 44309, 44310, 44311, 44312, 44313, 44314, 44315, 44316, 44317, 44318, 44319, 44320, 44321, 44322, 44323, 44324, 44325, 44326, 44327, 44328, 44329, 44330, 44331, 44332, 44333, 44334, 44335, 44336, 44337, 44338, 44339, 44340, 44341, 44342, |

| | |
|---|---|
| | 44343, 44344, 44345, 44346, 44602, 44603, 44604, 44605, 44606, 44607, 44608, 44609, 44610, 44611, 44612, 44613, 44614, 44615, 44616, 44617, 44618, 44619, 44620, 44621, 44622, 44623, 44624, 44625, 44626, 44627, 44628, 44629, 44630, 44631, 44632, 44633, 44634, 44635, 44636, 44637, 44638, 44639, 44640, 44641, 44642, 44643, 44644, 44645, 44646, 44647, 44648, 44649, 44650, 44651, 44652, 44653, 44654, 44655, 44656, 44657, 44658, 44659, 44660, 44661, 44700, 44701, 44799, 44800, 44820, 44850, 44899, 45000, 45005, 45020 |
| Rectum | 45110, 45111, 45112, 45113, 45114, 45115, 45116, 45117, 45118, 45119, 45120, 45121, 45122, 45123, 45124, 45125, 45126, 45127, 45128, 45129, 45136, 45150, 45395, 45397, 45800, 45805 |
| Anorectal | 45990, 46020, 46021, 46022, 46023, 46024, 46025, 46026, 46027, 46028, 46029, 46030, 46031, 46032, 46033, 46034, 46035, 46036, 46037, 46038, 46039, 46040, 46041, 46042, 46043, 46044, 46045, 46046, 46047, 46048, 46049, 46050, 46051, 46052, 46053, 46054, 46055, 46056, 46057, 46058, 46059, 46060, 46200, 46220, 46230, 46270, 46271, 46272, 46273, 46274, 46275, 46276, 46277, 46278, 46279, 46280, 46281, 46282, 46283, 46284, 46285, 46288 |

**Supplementary table 1: ICD and CPT codes**
